# Selumetinib in combination with dexamethasone for the treatment of relapsed/refractory RAS-pathway mutated paediatric and adult acute lymphoblastic leukaemia (SeluDex): study protocol for an international, parallel-group, dose-finding with expansion phase I/II trial

**DOI:** 10.1101/2021.10.22.21265327

**Authors:** Tobias Menne, Daniel Slade, Joshua Savage, Sarah Johnson, Julie Irving, Pamela R. Kearns, Ruth Plummer, Geoff Shenton, Gareth J. Veal, Britta Vormoor, Josef Vormoor, Lucinda Billingham

**Author notes:** Corresponding author contact details: Prof. Lucinda Billingham, Cancer Research UK Clinical Trials Unit (CRCTU), Institute of Cancer and Genomic Sciences, University of Birmingham, Edgbaston, Birmingham. B15 2TT. UK. These authors have contributed equally and should be considered joint first authors (Tobias Menne, Daniel Slade, Joshua Savage). These authors have contributed equally and should be considered joint senior authors (Josef Vormoor, Lucinda Billingham).

## Abstract

**Introduction:** Event free survival rates at 15 years for paediatric patients with relapsed/refractory acute lymphoblastic leukaemia (ALL) are 30-50%, with 5-year survival for adult patients only 20%. A large proportion of patients with newly diagnosed and relapsed ALL harbour somatic mutations that activate the RAS-signalling cascade. Steroids are a backbone of all induction blocks of ALL therapy, with preclinical data suggesting the combination of dexamethasone with the MEK1/2 inhibitor, selumetinib (ARRY-142886), results in a potent synergistic anti-cancer effect.

**Methods and analysis:** The SeluDex trial is an international, parallel-group, dose-finding with expansion, phase I/II trial to assess the selumetinib/dexamethasone combination in adult and paediatric patients with relapsed/refractory, RAS pathway mutant ALL. The Cancer Research UK Clinical Trials Unit at University of Birmingham is the UK Coordinating Centre, with national hubs in Copenhagen, Denmark; Monza, Italy; Münster, Germany; Paris, France; and Utrecht, Netherlands. Paediatric centres are all part of the Innovative Therapies for Children with Cancer consortium. Patients with morphologically proven relapsed/refractory or progressive B-cell precursor or T-ALL, with demonstrated RAS pathway activating mutations are eligible. Adult patients are ≥18 years old, ECOG ≤2 and paediatric <18 years old, Lansky play scale ≥60% or Karnofsky score ≥60%. The primary objective in phase I is to determine the recommended phase II dose of selumetinib as defined by occurrence/non-occurrence of dose limiting toxicities using the continual reassessment method, and phase II will evaluate preliminary anti-leukaemic activity of the selumetinib/dexamethasone combination, as defined by morphological response 28 days post treatment using a Bayesian approach. Target recruitment is between 26 and 42 patients (minimum of 13 and maximum of 21 in each group), depending on how many phase I patients are included also in phase II.

**Ethics and dissemination:** Medical ethical committees of all the participating countries will approve the study protocol. The results of this trial will be disseminated through national and international presentations and peer-reviewed publications.

**Trial registration number:** The trial was registered on EudraCT 2016-003904-29 on 21^st^ September 2016, ISRCTN 92323261, ClinicalTrials.Gov NCT03705507, and ITCC-063 study.

**Strengths and limitations of this study:**

- Novel combination of the MEK1/2 inhibitor, selumetinib, with dexamethasone
- Seamless phase I/II Bayesian trial design with a Continual Reassessment Method for dose escalation in phase I
- Parallel cohort trial design of adult and paediatric patients within one protocol
- Availability of CAR T-cell therapy since this trial started recruitment has competed for the same patient population
- Offers a bridging treatment option for patients awaiting CAR T-cell therapy outside clinical studies or at relapse after CAR-T treatment

## Introduction

Acute lymphoblastic leukaemia (ALL) is the most common childhood cancer, representing 26.8% of all childhood malignancies (1). While the overall cure rate for newly diagnosed paediatric ALL is approaching 90%, children with relapsed ALL (rALL) are still facing a poor outlook, with reported overall event-free survival rates at 15 years of 30-50% (2), and rALL remains a frequent cause of death.

In adults, the frequency of ALL is significantly lower, with an incidence of around one per 100,000 (3). ALL in adulthood has proven to be more challenging to treat compared to childhood ALL, with the disease being more resistant to chemotherapy, and patients have a reduced treatment tolerance especially in the elderly population. Five-year overall survival rate remains at ∼20% among adult patients aged ≥60 years, even though improvement has been observed since 1980 (4).

Data demonstrate that mutations activating the RAS-signalling cascade (NRAS, KRAS, FLT3 and PTPN11 and cCBL) are highly prevalent at both diagnosis and relapse of paediatric B-cell precursor ALL (incidence 38% at relapse), and are associated with high-risk features such as early relapse, central nervous system (CNS) involvement, and chemoresistance (5). Neurofibromin (NF1) copy number variants, point mutations or intragenic deletions have been reported in approximately 4% of T-cell ALL, 14% of early T-cell precursor (ETP), ∼30% of low hypodiploidy, 2% high risk of B-cell ALL and 75% of relapsed hypodiploid ALL (6-10). BRAF point mutation incidence is approximately 3% in T-cell ALL, 4% in ETP and 0.5% in high-risk B-cell ALL (7-9). IKZF2 and IKZF3 deletions which have recently been shown to activate the RAS-pathway have an incidence of approximately 18% in hypodiploid ALL (11). IL7Rα point mutations, deletions and in-frame alterations are found in approximately 9% of T-ALL (12, 13), and JAK1 point mutations are found in approximately 5% of T-ALL (14).

Selumetinib is a potent and selective allosteric MEK1/2 inhibitor that has been evaluated in phase II/III clinical trials for several cancers, including BRAF mutation-positive melanoma (15), metastatic uveal melanoma (16), pancreatic (17), colorectal (18), and KRAS mutation-positive NSCLC (19). Pre-clinical models suggest a synergistic effect when selumetinib and dexamethasone were combined in an orthotopic mouse model with three different RAS-pathway mutant ALL primagrafts and a mechanistic basis to the synergy was identified involving heightened levels of the proapoptotic Bim protein (20). However, the combination of selumetinib and dexamethasone has not been evaluated in a clinical trial setting.

The aim of the SeluDex trial is to determine the dose of selumetinib to use in combination with dexamethasone in adult and paediatric patient populations, and to assess the preliminary anti-leukaemic properties of the combination in each group of patients.

## Methods and analysis

### Study design

The SeluDex trial is a parallel-group, non-randomised, open-label, dose-finding with expansion, phase I/II clinical trial. As an early phase dose-finding and signal-seeking trial, a comparator arm was not included. Group A (adult patients) and Group P (paediatric patients) will both receive selumetinib in combination with a standard dose of dexamethasone (Figure 1). Patients will be recruited from public hospitals. The Cancer Research UK Clinical Trials Unit (CRCTU) at University of Birmingham is the UK Coordinating Centre, with national hubs in Copenhagen, Denmark; Monza, Italy; Münster, Germany; Paris, France; and Utrecht, Netherlands. The national coordinating and participating national centres are all part of the Innovative Therapies for Children with Cancer in Europe consortium (ITCC).

**Figure 1:**
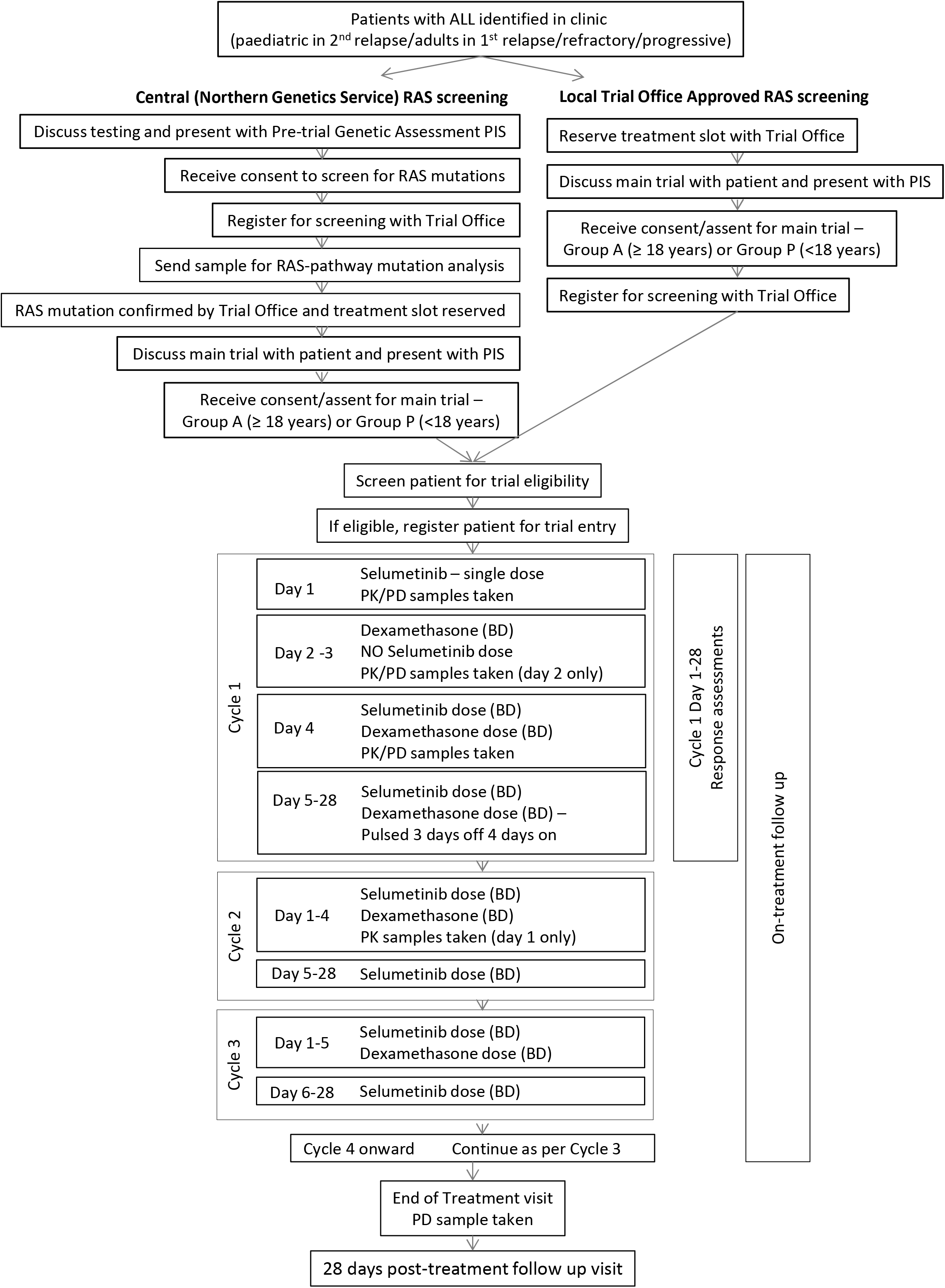
SeluDex trial schema. SeluDex trial schema showing the patient pathway highlighting screening, trial entry, treatment, and follow-up.

This trial aims to recruit between 26 and 42 patients (minimum of 13 and maximum of 21 in each group), depending on how many phase I patients are included also in phase II. The recruitment period is expected to be four years, with participants followed up for one month after a maximum of 6 treatment cycles. The World Health Organization (WHO) Trial Registration Data Set is attached in Supplementary Appendix 2.

### Patient and public involvement

Patients and public were not involved in the design and conduct of this research but will be consulted in the reporting and dissemination of its outputs.

### Patient selection

The two parallel patient groups in SeluDex, Group A (Adult) and P (Paediatric), have similar but discrete eligibility criteria. The main eligibility criteria are detailed in Tables 1 and 2, respectively. The screening process is the same for both groups. Eligibility for trial entry is dependent upon screening of a bone marrow aspirate to confirm morphological relapse and RAS-pathway mutation status. Peripheral blood may also be used in the case of a dry tap if white blood cell counts >50 × 10^9^/L. Centralised genetic analysis in the UK at Northern Genetics Service’s laboratories in Newcastle upon Tyne will identify the presence of a RAS-pathway activating mutation (key exons of NRAS, KRAS, FLT3, PTPN11 or cCBL mutation). A primary screen of the two most common mutations (NRAS and KRAS) will be performed first and if no mutations are found a secondary screen of the remaining mutations (FLT3, PTPN11 and cCBL) will be performed. Alternatively, locally obtained genetic results may be used to confer RAS-pathway mutation status if tested in a clinically compliant laboratory using a clinically validated test with material from the current relapse. Laboratories will be approved by the Trial Office to utilise local NRAS, KRAS, FLT3, PTPN11, cCBL, NF1, BRAF, IKZF2, IKZF3, IL7Rα or JAK1 testing.

**Table 1:**
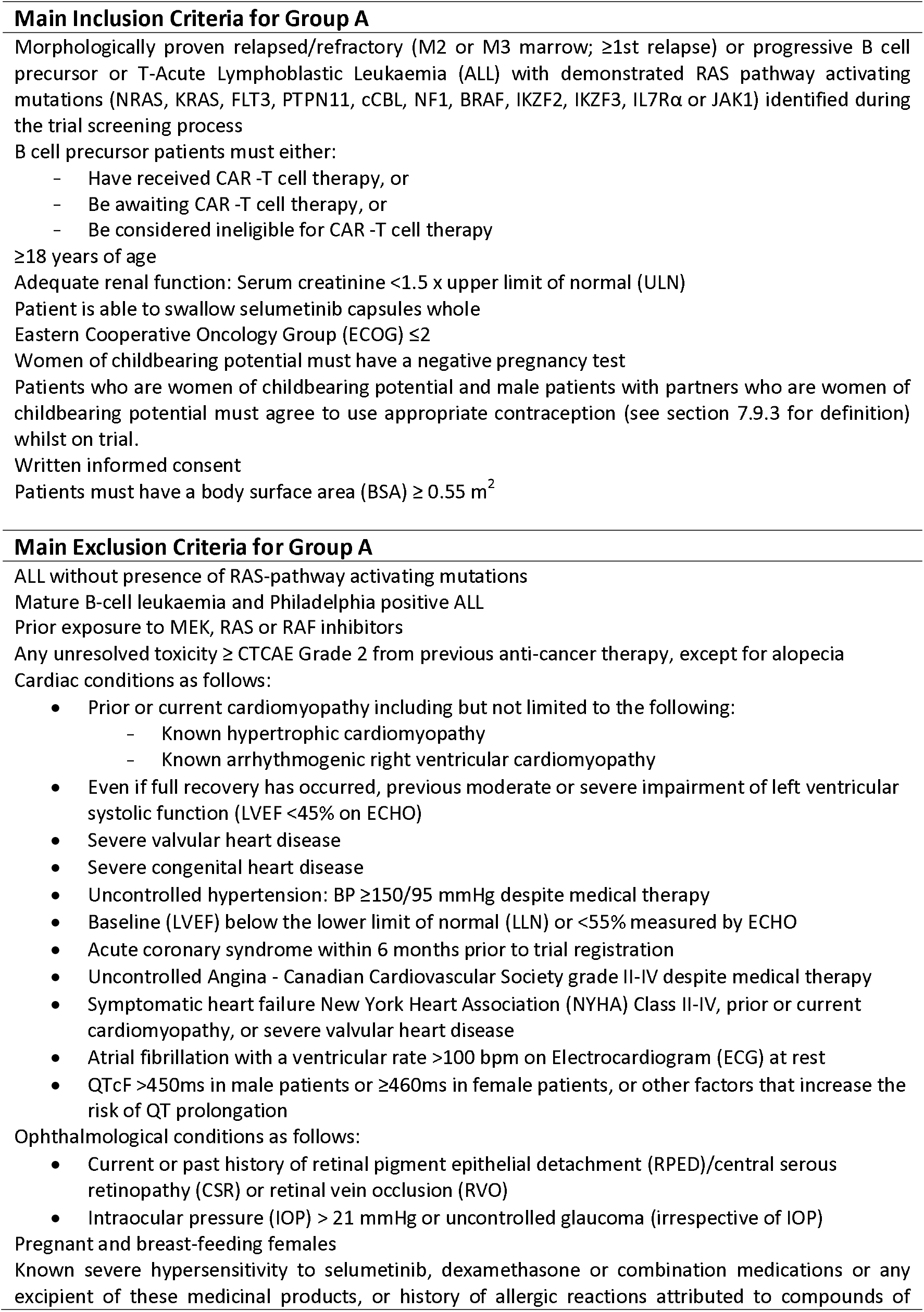
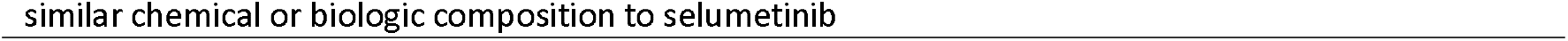
Main eligibility criteria for Group A in the SeluDex trial.

**Table 2:**
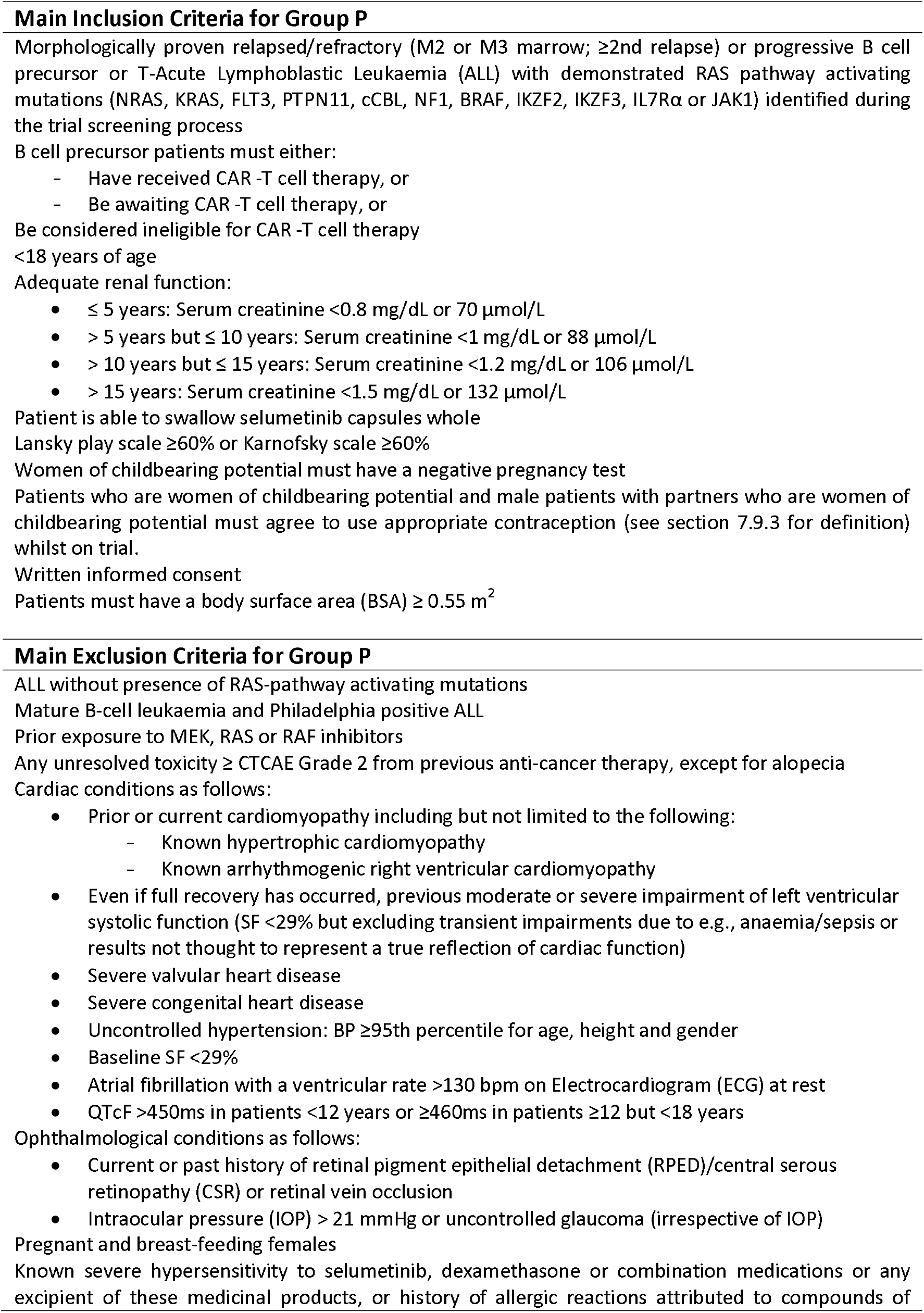
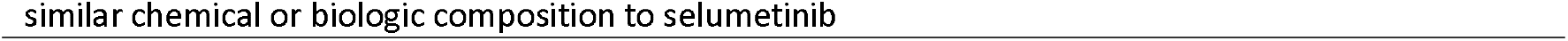
Main eligibility criteria for Group P in the SeluDex trial.

### Consent

Consent is requested by an investigator at two time points. A pre-trial genetic assessment consent allows for central/local analysis of a sample at screening to assess RAS mutation status, and the main trial consent, which is performed once mutation status is confirmed and a treatment slot is reserved. The main trial consent will be for either the phase I dose finding, of phase II dose expansion. Furthermore, age-specific consent/assent forms are available for patients over 16 years of age, and parents/legal guardians of paediatric patients (as appropriate according to age and national legislation). An example of the main consent form for adult patients is available in Supplementary Appendix 2. Age-specific patient information sheets are available for patients over 16 years of age, parents/legal guardians of paediatric patients, and teenage patients aged 13-15 years. As an example, the adult patients information sheet is available in Supplementary Appendix 3.

### Interventions

The dosing schedule for the dose-finding phase of the trial is summarised below for each group and detailed in relevant tables. Treatments are given over a 28-day cycle and will be continued for six cycles unless one or more of the following is observed; intolerable toxicity, confirmed disease recurrence or progression, pregnancy, severe non-compliance to protocol, development of any trial-specific criteria for discontinuation (not reaching at least a partial response (PR) at Cycle 1 day 28), or investigator decision, for example, if the patient requires a prohibited concomitant medication. The trial schedule of events is included in Supplementary Appendix 4.

#### Group A (Table 3)

**Table 3:** Group A (≥18 years) dosing schedule in phase I dose finding.

| **Dose Level** | **Selumetinib (% of adult MTD)** |  | **Dexamethasone** | **Initial Design**<br>Applicable prior to DLTs |
| --- | --- | --- | --- | --- |
|  | Cycle 1 D1: single dose<br>Cycle 1 D4-D28: BD dosing<br>Subsequent cycles 2-6: D1-28: BD dosing |  | Cycle 1 D2-4 & D8-11:<br>6mg/m<sup>2</sup>/day, D15-18 & D22-25: 4mg/m<sup>2</sup>/day<br>Cycle 2 D1-4: 4mg/m<sup>2</sup>/day<br>Subsequent cycles 3-6, D1-5 only 6mg/m<sup>2</sup>/day |  |
| -1 | 80% | 60mg BD PO | mg/m<sup>2</sup>*** /day PO divided into 2 doses (as per local practice) | n/a |
| 0 | 100% | 75mg BD PO |  | Cohorts 1 and 2 |
| +1* | 113-120%** | 85-90mg** BD PO |  | Cohorts 3, 4, 5 and 6 |

\* **Important:** The selumetinib dose reverts from the +1 dose level to dose level 0 with the start of cycle 2 as patients will only be exposed to a short pulse of dexamethasone which is not expected to substantially affect selumetinib PK
\*\* The proposed increase of dose might be changed based on PK results of dose level 0
\*\*\* Body surface area (BSA) for Group A to be calculated according to standard institutional practice

##### Dexamethasone

Steroids are a backbone of all induction blocks of ALL therapy with dexamethasone, the preferred steroid in the management of patients with ALL. Initially, dexamethasone was to be administered on a continuous dosing schedule with a weekly total dose of 42mg/m^2^ for the first course. However, following an observed increase in the risk of infections during the initial stages of the trial, the trial dexamethasone dosing regimen was amended to allow intermittent treatment in blocks rather than continuously, and incorporating a dose reduction from 6mg/m^2^/day in cycle 1 (D2-4 & D8-11) to 4mg/m^2^/day during D15-18 and D22-25 of cycle 1. Further dexamethasone is given as a low dose in cycle 2 (D1-4 only: 4mg/m2/day) and then as a high dose again in subsequent cycles (C3-6, D1-5 only: 6mg/m^2^/day).

##### Selumetinib

The dose of 75mg was chosen as the starting dose for Group A (Dose Level 0) with the dose only increased if pharmacokinetic (PK) data suggest the addition of dexamethasone decreases selumetinib serum concentrations significantly due to induction of the cytochrome P450 enzyme CYP3A4 (21).

#### Group P (Table 4)

**Table 4:** Group P (<18 years) dosing schedule in phase I dose finding.

| **Dose Level** | **Selumetinib (% of adult MTD)** |  | **Dexamethasone** | **Initial Design**<br>Applicable prior to DLTs |
| --- | --- | --- | --- | --- |
|  | Cycle 1 D1: single dose<br>Cycle 1 D4-D28: BD dosing<br>Subsequent cycles 2-6: D1-28: BD dosing |  | Cycle 1 D2-4 & D8-11:<br>6mg/m<sup>2</sup>/day, D15-18 & D22-25: 4mg/m<sup>2</sup>/day<br>Cycle 2 D1-4: 4mg/m<sup>2</sup>/day<br>Subsequent cycles 3-6, D1-5 only 6mg/m<sup>2</sup>/day |  |
| -1 | 80% | 20mg/m<sup>2***</sup> BD PO | mg/m<sup>2***</sup>/day PO divided into 2 doses (as per local practice) | Cohorts 1 and 2 |
| 0 | 100% | 25mg/m<sup>2***</sup> BD PO |  | Cohorts 3 and 4 |
| +1* | 113-120%** | 28-30mg<sup>**</sup>/m<sup>2***</sup> BD PO |  | Cohorts 5 and 6 |

\* **Important:** The selumetinib dose reverts from the +1 dose level to dose level 0 with the start of cycle 2 as patients will only be exposed to a short pulse of dexamethasone which is not expected to substantially affect selumetinib PK
\*\* The proposed increase of dose might be changed based on PK results of dose level 0
\*\*\* BSA for Group P to be calculated according to standard institutional practice

##### Dexamethasone

As with adult dosing, trial dexamethasone dosing regimen was amended during the initial stages of the trial to be reduced after the first two weeks and to be given pulsed during cycle 1 due to the increase in infection risk.

##### Selumetinib

Results of two paediatric phase I trials using selumetinib only (22, 23) gave the maximum tolerated dose (MTD) as 2 × 25mg/m^2^; thus in the SeluDex trial, 20mg/m^2^ (80% of the single agent MTD of selumetinib) was chosen as the starting dose for safety reasons (Dose Level -1), specifically for those patients with CNS disease receiving concomitant intrathecal methotrexate.

### Treatment compliance

Treatment compliance should be discussed with the patient/parent/legal guardian at the beginning of each cycle when study drug is dispensed. The research nurse should complete the relevant sections including recording the morning and afternoon doses and the dates they are to be taken on the diary. All patients/parents/legal guardians will be required to complete a diary with the daily time of administration of the study drugs, which must be returned to the clinic for checking at each visit. If a dose is missed, the reason must be noted in the diary by the patient/parent/legal guardian. Patients/parents/legal guardians should be advised to return any unused investigational medcicinal product in the original bottles, in addition to returning any empty bottles. Compliance should be reviewed at the end of each cycle by the site staff via review of the patient diary and discussion with the patient/parent/legal guardian that is to be documented in the medical notes.

### Dose modifications

No dexamethasone dose reductions are permitted during cycle 1 and 2. If identifiable dexamethasone related intolerable toxicity is present during cycle 3 then a dose reduction to 4mg/m^2^/day is allowed from cycle 4 onwards.

Dose modifications for the following selumetinib-related toxicities are allowed:

- Grade 3 neutropenia with infection and/or fever (GCSF is permitted)
- Grade 3 or 4 nausea, vomiting, or diarrhoea, if persistent despite optimal antiemetic and/or anti-diarrhoeal therapy
- Any Grade 3 or greater non-haematological toxicity
- Any other Grade 4 haematological toxicity

If one or more of these are observed the following action should be taken;

1^st^ occurrence: Hold selumetinib until recovery to Grade <1 or baseline; restart at original dose level

2^nd^ occurrence: Hold selumetinib until recovery to Grade <1 or baseline; keep same dose but reduce to once daily

3^rd^ occurrence: Hold selumetinib until recovery to Grade <1 or baseline; use 60% of the original dose but go back to twice daily. For group P: 45mg reduce to 30 mg; 40 mg to 25 mg; 30 – 35 mg to 20 mg; 20 – 25 mg to 10 mg. For Group A: 75 mg reduce to 45 mg; 60 mg to 35 mg

4^th^ occurrence: Discontinue selumetinib

### Concomitant medication

Patients should avoid consuming large amounts of grapefruits, seville oranges, and any other products that may contain these fruits, e.g., grapefruit juice. In addition, patients should not take any vitamin E supplements. Restricted medications are listed in in Table 5.

**Table 5:** Restricted medications within the SeluDex trial.

|  |  |  |
| --- | --- | --- |
| • Acetazolamide<br>• Aminoglutethimide<br>• Antacids<br>• Anticholinesterases<br>• Anti-hypertensives<br>• Aprepitant<br>• Barbiturates<br>• Beta-naphthoflavone<br>• Carbamazepine<br>• Carbenoxolone<br>• Clarithromycin<br>• Coumarin anticoagulants<br>• Diltiazem<br>• Diuretics<br>• Efavirenz<br>• Ephedrine<br>• Erythromycin | • Fluconazole<br>• Fluvoxamine<br>• Glucocorticoids<br>• Hypoglycaemic agents, e.g. insulin<br>• Indinavir<br>• Itraconazole<br>• Ketoconazole<br>• Loop diuretics<br>• Methylcholanthrene<br>• Modafinil<br>• Nafcillin<br>• Nefazodone<br>• Nelfinavir<br>• Nevirapine<br>• Norethindrone<br>• NSAIDs<br>• Omeprazole | • Oral contraceptives<br>• Oxcarbazepine<br>• Phenobarbital<br>• Phenytoin<br>• Pioglitazone<br>• Prednisolone<br>• Primidone<br>• Rifabutin<br>• Rifampicin<br>• Ritonavir<br>• Salicylates<br>• Saquinavir<br>• Suboxone<br>• Telithromycin<br>• Thiazide diuretics<br>• Troglitazone<br>• Verapamil<br>• St John's Wort |

### Trial outcomes

#### Phase I

The primary objective for the phase I dose-finding component is to determine the recommended phase II dose (RP2D) of selumetinib in combination with dexamethasone, as defined by the occurrence/non-occurrence of dose limiting toxicities (DLTs) in the trial-defined assessment period. The trial will achieve this through the use of the continual reassessment method (CRM) (24). DLTs will be assessed from the first dose on cycle 1, day 1 up until cycle 1, day 28 during phase I only. A DLT is defined as any toxicity which is dose limiting, is not attributable to the disease or disease-related processes under investigation, and is considered at least possibly related to selumetinib or dexamethasone (see Dose Limiting Toxicity section).

Secondary outcomes for the phase I component are:

- The occurrence of adverse events (AEs) as measured by Common Terminology Criteria for Adverse Events (CTCAE) version 4 (25) and causality assessment
- Pharmacokinetic variables of selumetinib in combination with dexamethasone from the concentration time profile (area under the plasma concentration-time curve (AUC), C_max_, T_max_, t_1/2_)
- Response to treatment assessed by complete remission rate at 28 days as measured by morphological and minimal residual disease (MRD) response in bone marrow (BM) and for patients with CNS involvement only clearance of cerebrospinal fluid (CSF) blasts at 28 days
- Difference in pharmacokinetics of selumetinib (ΔAUC) when selumetinib is administered as single agent and in combination with dexamethasone

#### Phase II

The primary objective for the phase II expansion component will be to assess preliminary anti-leukaemic activity of the selumetinib/dexamethasone drug combination as defined by morphological response (complete remission (CR), complete remission with incomplete platelet recovery (CRi), partial remission (PR), non-response (NR) and for patients with CNS involvement only, clearance of CSF blasts) 28 days post-treatment end. Response evaluation will be undertaken with bone marrow response at day 28, including MRD and CSF surveillance for CNS positive patients with standard cytology. A patient will be classified as having a response if they achieve CR or CRi at the day 28 assessment (with an acceptable window of day 15 to day 35).

Secondary outcomes for the phase II component are;

- The occurrence of AEs as measured by CTCAE version 4 and causality assessment
- The occurrence/non-occurrence of DLTs in the trial defined assessment period
- Pharmacokinetic variables of selumetinib in combination with dexamethasone from the concentration time profile (AUC, C_max_, T_max_, t_1/2_)
- Difference in pharmacokinetics of selumetinib (ΔAUC) when selumetinib is administered as single agent and in combination with dexamethasone
- MRD response in bone marrow at 28 days

Exploratory outcomes of the trial for both phases of the trial include exploratory pharmacodynamic (PD) biomarker studies if clinical responses are observed. These will include levels of phosphorylated ERK by flow cytometry in leukaemic cells from patients at several time points (cycle 1 day 1, cycle 1 day 4 and end of treatment visit), as well as retrospective mRNA profiling, including the apoptotic initiator BIM (20, 26).

### Dose limiting toxicities

DLTs will be assessed from the first dose on cycle 1 day 1 up until cycle 1 day 28 during Phase I. A haematological DLT is defined as bone marrow aplasia/hypoplasia, defined as overall marrow cellularity less than 25% (malignant infiltration or other causes are excluded), which does not recover by cycle 1 day 28. This includes an absolute neutrophil count of less than 0.5×10^9^/L and a non-transfusion dependent platelet count of less than 25×10^9^/L due to bone marrow aplasia/hypoplasia. In addition, Grade 4 febrile neutropenia, and any Grade 5 AE are also defined as a haematological DLT.

Non-haematological DLTs in Group A are defined as Left Ventricular Ejection Fraction <45% as measured by echocardiogram; and in Group P as >15% reduction in Shortening Fraction from baseline by echocardiogram. Results must be viewed by a consultant cardiologist to confirm that there has been a genuine deterioration in cardiac function. In addition, any Grade 5 AE, Grade 3 nausea or vomiting on or before C1D28 only which does not resolve to <Grade 2 within 48 hours (with or without supportive care), Grade 3 mucositis on or before C1D28 only which does not resolve to <Grade 2 within 48 hours (with or without supportive care), any other Grade 3 or 4 non-haematological toxicity, specifically excluding: Alopecia; dexamethasone related CTCAE Grade 3 toxicities (e.g., hyperglycaemia, muscle weakness, psychosis, avascular necrosis); tumour lysis syndrome; and increased alanine transaminase or aspartate aminotransferase.

Suspected DLTs will be reviewed and confirmed by the TSC during dose decision meetings at the end of each cohort during Phase I.

### Statistical analysis plan

Both groups (A and P) will be analysed separately for both phases of the trial. If the response rates in the two groups are seen to be similar in the phase II component of the trial, then there may be some consideration to analysing response in the adult and paediatric groups collectively. The phase I evaluable population includes those patients who do not withdraw/die or discontinue treatment due to non-treatment related causes before the end of the DLT assessment period. A patient is evaluable regardless of the above if they experience a DLT. The phase II evaluable population includes all patients who received any treatment. There is no scope to replace unevaluable patients in the trial.

#### Phase I

The maximum number of patients to be recruited in phase I is 24; 12 into each group in parallel.

The trial will utilise the continual reassessment method (CRM) to find which doses to take forward to the phase II activity component of the trial for each group respectively. Each group will use a modified two-stage Bayesian CRM (27), recruiting patients in cohorts of two. The design incorporates rules to enable early stopping if the DLT rate at the lowest dose is unacceptably high. Patients are assigned to the starting dose level and follow the initial design (see Tables 3 and 4) until the first DLT is observed. Once the first DLT is observed the CRM takes over and determines the ‘best’ dose level that the next cohort should be assigned to, based on the model-based predicted probabilities of toxicity at each possible dose. The acceptable level of toxicity (probability of a DLT) has been chosen to be 17% for each group, that is, the trial will aim to find the highest dose that coincides with this acceptable level. This value was selected based on discussion with clinicians concerning the acceptable rate of DLTs based on the chosen definition.

The stopping rule for toxicity utilised in the modified CRM is based on the probability of the true toxicity rate at the lowest dose being >27%. If based on the observed data the probability of the true rate of toxicity being greater than this chosen value is >0.85 then the modified CRM will suggest that the trial should stop. The current MTD estimate is calculated after each cohort has been assessed for the full DLT assessment period and the next cohort of patients are then assigned to this MTD estimate. The final MTD estimate calculated after all 12 patients have been treated is the dose estimate to be considered to be taken forward to the phase II component.

This design works relatively well with computer simulations showing a probability for correctly selecting the right dose level of at least 0.6 when it is truly dose level 0 or 1, around 0.5 when it is truly dose level - 1 and at least 0.6 for stopping early when all doses are truly too toxic.

Data will be reviewed at each dose decision stage by the independent Trial Safety Committee (TSC). The TSC will incorporate PK results into the decision process; the CRM will advise as to which doses are safe, with the PK additionally informing if there is a need for escalation/de-escalation.

#### Phase II

The phase II part of the trial will include patients from the phase I component that were dosed at the RP2D plus patients recruited into the expansion cohort. The number of phase I patients taken forward to the phase II assessment will be at least 4 in each group and the maximum number of patients to be recruited in the expansion cohort will be 9 in each group, giving a total of 13.

Based on data published by the TACL group (CR rate of 44% for second relapses and 27% for third relapses (28)) and seeing as we expect a mixture of 2^nd^, 3^rd^ and higher relapses in paediatric patients and similar if not worse relapse rates for adult patients, we estimate the overall response rate on standard treatment to be approximately 35%.

This phase will utilise a Bayesian approach to investigate the true response rate in each group. A Beta-Binomial conjugate analysis will be used to create a posterior probability distribution of the true response rate using the observed trial data combined with a Beta (1,1) minimally informative prior.The trial design is based on the decision criteria that if there is a high probability (>0.80) that the true response rate is >35% then this will indicate that the treatment is worthy of further research. With 13 patients, this design works reasonably well with a low probability of 0.08 of incorrectly concluding that the treatment is worthy when the true response rate is only 25% and a high probability of 0.82 of correctly concluding that the treatment is worthy when the true response rate is 55%.

If the response rates are similar in both groups, then information may be borrowed across Groups A and P to provide more accuracy in determining the true response rate for both groups collectively

There are no planned subgroup or interim analyses.

### Pharmacokinetic and pharmacodynamic samples

Blood samples for PK and PD analysis will be obtained on days one, two and four of cycle 1, with blood samples for PK analysis collected again on day one of cycle 2, and blood samples for PD analysis obtained at the end of trial treatment.

For PK analysis plasma will be subjected to liquid chromatography-mass spectrometry (LC-MS) analysis to quantification concentrations of selumetinib, its major metabolite N-desmethyl selumetinib and dexamethasone. Two types of PD samples will be prepared from peripheral blood samples of patients who have a white cell count greater than 10×10^9^/L; samples will either be fixed on site in BD Lyse Fix solution or collected separately in Blood RNA Preservative Tubes. These PD samples will be transferred to the UK Biobank at the University of Birmingham for storage.

All samples will be collected in accordance with national regulations and requirements including standard operating procedures for logistics and infrastructure. Samples will be taken in appropriately licensed premises, stored and transported in accordance with the Human Tissue Authority guidelines and NHS trust policies.

### Adverse events reporting and analysis

The collection and reporting of AEs as measured by CTCAE v4 will be in accordance with the Research Governance Framework for Health and Social Care and the requirements of the National Research Ethics Service (NRES). Definitions of different types of AEs are listed in online Supplementary Appendix 5; this includes abnormal laboratory findings which are considered clinically significant. The reporting period for AEs will be from the date of commencement of protocol defined treatment until 28 days after the administration of the last treatment. The investigator should assess the seriousness and causality (relatedness) of all AEs experienced by the patient (this should be documented in the source data) with reference to the protocol. In addition, DLTs will be assessed for the Phase I part of the trial only. All will be reported using the applicable electronic case report form (eCRF).

### Data management

Case report forms (CRFs) can be entered online at https://www.cancertrials.bham.ac.uk/SeluDexLive. Authorised staff at sites will require an individual secure login username and password to access this online data entry system. Paper CRFs must be completed, signed/dated and returned to the SeluDex Trial Office by the investigator or an authorised member of the site research team. Data reported on each CRF should be consistent with the source data or the discrepancies should be explained. If information is not known, this must be clearly indicated on the CRF. All missing and ambiguous data will be queried. All sections are to be completed.

All trial records must be archived and securely retained for at least 25 years. No documents will be destroyed without prior approval from the sponsor, via the SeluDex Trial Office. On-site monitoring will be carried out as required following a risk assessment and as documented in the Quality Management Plan. Any monitoring activities will be reported to the central SeluDex Trial Office and any issues noted will be followed up to resolution. SeluDex will also be centrally monitored, which may trigger additional on-site monitoring. Further information regarding data management is provided in the study protocol.

The CRCTU will hold the final trial dataset and will be responsible for the controlled sharing of anonymised clinical trial data with the wider research community to maximise potential patient benefit while protecting the privacy and confidentiality of trial participants. Data anonymised in compliance with the Information Commissioners Office requirements, using a procedure based on guidelines from the Medical Research Council (MRC) Methodology Hubs, will be available for sharing with researchers outside of the trials team within 12 months of the primary publication.

### Trial organisation structure

The University of Birmingham will act as sponsor to this international, multi-centre study: Support Group, Aston Webb Building, Room 119, Birmingham, B15 2TT. The trial is being conducted under the auspices of the CRCTU, The University of Birmingham according to their local procedures. The Trial Management Group (TMG) will be responsible for the day-to-day running and management of the trial. Members of the TMG include the chief investigator, coinvestigators, biological coordinators, trial management team leader, senior trial coordinator, trial coordinator, lead trial statistician and trial statistician. The TMG will have regular meetings during recruitment. The TSC will consist of independent clinicians, Dr Bela Wrench and Dr Christina Halsey, as well as an independent statistician, Dr Graham Wheeler.

Data analyses will be supplied in confidence to the independent TSC, which will be asked to support dose decisions and advise on whether the accumulated data from the trial, together with the results from other relevant research, justify the continuing recruitment of further patients. The TSC will operate in accordance with a trial specific charter based on the template created by the Damocles Group. The TSC will meet at the end of every cohort in phase I and at least every 6 months in phase II, or more often if required e.g., an emergency meeting may also be convened if a safety issue is identified. The TSC will report directly to both the SeluDex TMG who will convey the findings of the TSC to the funders/sponsor as appropriate or when specifically requested by these parties.

### Confidentiality

Confidential information collected during the trial will be stored in accordance with the General Data Protection Regulation (GDPR) 2018. As specified in the patient information sheet (PIS) and with the patient’s consent, patients will be identified using only their initials, date of birth and unique trial ID number. Authorised staff may have access to the records for quality assurance and audit purposes. The Trials Office maintains the confidentiality of all patients’ data and will not disclose information by which patients may be identified to any third party other than those directly involved in the treatment of the patient and organisations for which the patient has given explicit consent for data transfer (e.g., laboratory staff).

### Trial status

Recruitment for the trial opened in April 2018 and recruitment is expected to last until December 2021. A list of open sites can be obtained from the SeluDex Trial Office.

## Discussion

### Position of the trial in an era of CAR-T cell treatment

The SeluDex trial was initially proposed in 2013, before the advent of CAR-T cell and other CD19 and CD22 targeting immunotherapies. This trial was designed for patients who had relapsed after stem cell transplantation, with little or no other therapeutic options. The trial opened in the UK in Spring 2018. Since commercial CAR-T cell treatment (tisagenlecleucel) was approved by the FDA/EMA in 2018, eligible patients were preferentially offered CAR-T cell treatment over enrolment in this experimental study, and this has led to a significant drop in expected trial recruitment. After two years of experience with tisagenlecleucel, relapse after cellular immunotherapy is emerging as a new therapeutic challenge (29). Relapses after CAR-T cell failure, especially CD19 negative relapses, and relapsed/refractory T cell ALL should be considered/screened for the SeluDex trial. For patients who have not had CD19 or CD22 targeting immunotherapy these should be prioritised, however, in exceptional circumstances the SeluDex trial (e.g., cycle 1) may also be considered as bridging therapy before CAR-T cell infusion.

### Implemented urgent safety measures

Since the study opened to recruitment in 2018 there have been three urgent safety measures (USM) implemented in the protocol to date. The first USM was in December 2018 to reduce the dose of dexamethasone after two weeks during cycle 1 for Group A patients, after an adult patient experienced a fatal Suspected Unexpected Adverse Reaction (SUSAR) from rapid onset gram-negative sepsis; investigators suspected that the steroid administration had masked potential symptoms of infection and sepsis. The second USM was implemented in May 2019 to reduce the dose of dexamethasone after two weeks during cycle 1 for Group P patients, in line with the measure already implemented for adults, in addition to mandating fluoroquinolone prophylaxis during cycle 1 for all patients. This was due to a further fatal SUSAR of gram-negative sepsis in a paediatric patient. The third USM implemented in September 2019 changed the dexamethasone dose to pulsed during cycle 1 and mandated fluoroquinolone prophylaxis in cycle 2, in addition to cycle 1, and co-trimoxazole prophylaxis throughout trial treatment for all patients. This was due to a further fatal SUSAR in an adult patient from pneumonia considered related to trial treatment. Pulsed dosing of dexamethasone (four days on, three days off) is standard practice in the management of adult ALL patients and is better tolerated than continuous dexamethasone dosing during induction. Since these USMs were implemented, there have been no further events of concern and the TMG, TSC and regulatory authorities are confident that the potential patient benefit outweighs the risks of the intervention.

### Trial strengths

This trial is, to the best of our knowledge, one of the first to span the whole age range of paediatric and adult patients. On the paediatric side, it is only limited because participants need to swallow the selumetinib capsules whole. As soon as a paediatric liquid formulation is available, all age limitations could be removed.

The use of the CRM greatly also aided in the delivery of this trial, providing the flexibility to utilise cohorts of only two patients allowing for more frequent dose adjustment. Additionally, the CRM provides the means to estimate probabilities of DLT at all doses along with associated uncertainty intervals for those estimates, even those untested doses. The CRM in general identifies the MTD with greater accuracy and treats more patients at the MTD than traditional rule-based methods e.g. 3+3 (30).

Finally, the paediatric part of the SeluDex trial is an approved Innovative Therapies for Children with Cancer in Europe (ITCC) study with five National Co-ordinating Centres (NCC) outside the UK. The NCC of the Netherlands opened in Summer 2020, with Germany, France, Italy and Denmark planned to follow in 2021. This will help to improve the slow recruitment numbers resulting from the introduction of CAR-T cell therapy.

### Ethics and dissemination

The trial will be performed in accordance with the recommendations guiding physicians in biomedical research involving human subjects, adopted by the 18^th^ World Medical Association General Assembly, Helsinki, Finland and stated in the respective participating countries laws governing human research, and Good Clinical Practice. The protocol was approved by the medical ethical committees of all the participating countries. UK ethics approval (17/YH/0123) was granted by Yorkshire & The Humber – Leeds West Research Ethics Committee on 12^th^ July 2017. Initial UK Competent Authority approval was granted by Medicines and Healthcare Products Regulatory Agency (MHRA) on 5^th^ May 2017, with subsequent protocol versions approved; current version in use is 9.0, 22^nd^ March 2021.

A meeting will be held after the end of the study to allow discussion of the main results among the collaborators before publication. Results of the primary and secondary endpoints will be submitted for publication in peer-reviewed journals. Manuscripts will be prepared by the TMG and authorship determined by mutual agreement.

### Summary

In summary, the SeluDex trial investigates the novel combination of the MEK1/2 inhibitor, selumetinib, with dexamethasone in a seamless phase I/II Bayesian trial design, in both adult and paediatric relapsed/refractory ALL patients. Although CAR T-cell therapy was approved after the trial started recruitment, the trial may serve as a bridging treatment option for patients awaiting CAR T-cell therapy and a treatment option for relapses failure once CAR-T cell treatment has failed.

## Supporting information

SPIRIT

## Data Availability

Participant data and the associated supporting documentation will be available within 6 months after the publication of this manuscript. Details of our data request process is available on the CRCTU website. Only scientifically sound proposals from appropriately qualified research groups will be considered for data sharing. The decision to release data will be made by the CRCTU Director's Committee, who will consider the scientific validity of the request, the qualifications and resources of the research group, the views of the Chief Investigator and the trial steering committee, consent arrangements, the practicality of anonymising the requested data and contractual obligations. A data sharing agreement will cover the terms and conditions of the release of trial data and will include publication requirements, authorship and acknowledgements and obligations for the responsible use of data. An anonymised encrypted dataset will be transferred directly using a secure method and in accordance with the University of Birmingham's IT guidance on encryption of data sets.

## Contributors

TM: Chief Investigator, study conception, study design, protocol development. DS: Senior Trial Biostatistician, study design, statistical plan development. JS: Trial Team Leader, study management, protocol development, coordination of IRAS, REC, MHRA, HRA and local R&D applications. SJ: Trial Coordinator, protocol development, coordination of IRAS, REC, MHRA, HRA and local R&D applications. PRK: Co-Investigator, study conception, study design, protocol development. JV: International Lead Investigator, study conception, study design, protocol development. BV: Co-Investigator, study conception, study design, protocol development. JI: Biological Coordinator, study conception, study design, protocol development. GV: Biological Coordinator, study conception, study design, protocol development. GS: Co-Investigator, protocol development. RP: Co-Investigator, study conception, study design, protocol development. LB: Chief Biostatistician, study conception, study design, protocol development, statistical plan development.

## Acknowledgements

The Investigators and Sponsor thank all the patients and their families who participated in this trial, as well as the NHS Trusts and staff and the members of the Trial Safety Committee, chaired by Dr Bela Wrench, who have supported this trial. SeluDex was supported by Experimental Cancer Medicine Centres (ECMC) funding and by the ECMC Network. This trial has been independently peer reviewed and has been adopted by the National Institute for Health Research Clinical Research Network Portfolio. Special thanks to Dr Siân Lax for her contributions to this paper.

