## Supplementary Appendix 1 for "Selumetinib in combination with dexamethasone for the treatment of relapsed/refractory RAS-pathway mutated paediatric and adult acute lymphoblastic leukaemia (SeluDex): study protocol for an international, parallel-group, dose-finding with expansion phase I/II trial"

| Data category | Information |
|---|---|
| Primary registry and trial identifying number | EudraCT Number<br>2016-003904-29 |
| Date of registration in primary registry | 21-Sep-2021 |
| Secondary identifying numbers | ISRCTN: 92323261<br>ClinicalTrials.Gov: NCT03705507<br>ITCC-063 study |
| Source(s) of monetary or material support | Cancer Research UK<br>AstraZeneca<br>National Coordinating Centres were supported by a grant from ITCC "Imagine for Margo" fund and AstraZeneca |
| Primary sponsor | University of Birmingham |
| Secondary sponsor(s) | n/a |
| Contact for public queries | LB: |
| Contact for scientific queries | TM: |
| Public title | A trial looking at selumetinib and dexamethasone for acute lymphoblastic leukaemia (SeluDex) |
| Scientific title | SeluDex: an international trial of selumetinib in combination with dexamethasone for the treatment of acute lymphoblastic leukaemia |

| **Data category** | **Information** |
|---|---|
| Countries of recruitment | UK, Denmark, Italy, Germany, France, Netherlands |
| Health condition(s) or problem(s) studied | Relapsed/refractory acute lymphoblastic leukaemia |
| Intervention(s) | Dexamethasone and selumetinib |
| Key inclusion and exclusion criteria: **Adult Group** | Ages eligible for study: ≥18 years<br>Sexes eligible for study: both<br>Accepts healthy volunteers: no |
| | Inclusion criteria: adult patient (≥18 years) with proven ALL with demonstrated RAS pathway activating mutations, performance status ≤2 |
| | Exclusion criteria: Prior exposure to MEK, RAS or RAF inhibitors, pregnancy or breastfeeding females, cardiac and/or ophthalmology conditions |
| Key inclusion and exclusion criteria: **Paediatric Group** | Ages eligible for study: <18 years<br>Sexes eligible for study: both<br>Accepts healthy volunteers: no |
| | Inclusion criteria: paediatric patient (<18 years) with proven ALL with demonstrated RAS pathway activating mutations, able to swallow selumetinib capsules whole, Lansky play scale ≥60% or Karnofsky scale ≥60% |
| | Exclusion criteria: Prior exposure to MEK, RAS or RAF inhibitors, pregnancy or breastfeeding females, cardiac and/or ophthalmology conditions |
| Study type | Interventional |
| | Allocation: non-randomised, open-label |

| Data category | Information |
|---|---|
| | Primary purpose: dose-finding and preliminary efficacy |
| | Phase I/II |
| Date of first enrolment | 18-May-2018 |
| Target sample size | Between 26 and 42 patients; minimum of 13 and maximum of 21 in each group, |
| Recruitment status | Open |
| Primary outcome(s) | Phase I: Occurrence/non-occurrence of dose limiting toxicities (time frame: between days 1 and 28 of cycle 1)<br>Phase II: Morphological response (time from: 28 days post-treatment end) |
| Key secondary outcome(s) | Occurrence of AEs as measured by CTCAE version 4 and causality assessment<br>Pharmacokinetic variables of selumetinib in combination with dexamethasone from the concentration time profile |
