## Supplementary Appendix 2 for "Selumetinib in combination with dexamethasone for the treatment of relapsed/refractory RAS-pathway mutated paediatric and adult acute lymphoblastic leukaemia (SeluDex): study protocol for an international, parallel-group, dose-finding with expansion phase I/II trial"

***Please delete this line and print on hospital headed paper***

### Informed Consent Form – Adult, Phase I

*To be used only for participants over 16 years of age entering dose finding phase*

EudraCT Reference: 2016-003904-29

Site: ______________________________

Principal Investigator: ______________________________

Trial Number (TNO): ☐☐☐ Screening Number (SNO): ☐☐ / ☐☐

| | Please INITIAL each box |
|---|---|
| 1. I confirm that I have read and understand the ***Participant Information Sheet – Adult, Phase I*** (version…….. dated…………………………) for the above trial. I have had the opportunity to consider the information, ask questions, and have had these answered satisfactorily | ☐ |
| 2. I understand that my participation is voluntary and that I am free to withdraw at any time without giving any reason, without my medical care or legal rights being affected. I understand that if I withdraw from treatment my doctor may continue to provide the Trial Office with information that would routinely be collected about me and recorded in my medical notes. I am aware that I can also withdraw consent for this data transfer | ☐ |
| 3. I give permission for my initials, date of birth, hospital number and NHS number to be given to the SeluDex Trial Office when I am registered to the trial as well as a copy of this consent form | ☐ |
| 4. I understand that relevant sections of my medical notes and data collected during the trial may be looked at by individuals from the SeluDex Trial Office, regulatory authorities, Sponsor and/or NHS bodies, where it is relevant to my taking part in this research. I understand that this information will be held in a confidential manner. I give permission for | ☐ |

these individuals to have access to my records.

5. I understand that anonymised data from the trial may be provided to other third parties (e.g. pharmaceutical companies or other academic institutions) for research, safety monitoring or licensing purposes

6. I agree to my GP being informed of my participation in this trial

7. I understand that the SeluDex Trial Office, may access information held by national cancer registries and within national databases to keep in touch with me and to follow up on my health status

8. I give permission for collection of samples of my blood and bone marrow to be used in the SeluDex trial. I understand that samples will be sent to the University of Birmingham and other laboratories in the United Kingdom

9. I understand that DNA analysis may be performed on the samples taken for the trial.

10. I agree to take part in the above trial

**The following is optional and will not affect entry into the trial, please initial in the relevant box:**

| | No | Yes |
|---|---|---|
| If in the event that an abnormality that might affect other family members is uncovered during genetic testing for the trial, I would like my doctor to be informed and to be referred to a genetic counsellor if appropriate. | | |

**Name of participant** **Date** **Signature**

**Name of person taking consent** **Date** **Signature**

You must have signed the Site Signature & Delegation Log

This document was drafted using CRCTU-ICF-QCD-001, version 2.0

Original to be kept in the Investigator Site File, 1 copy in hospital notes, 1 copy to the participant, 1 copy to the National Coordinating Centre
