## Supplementary Appendix 3 for "Selumetinib in combination with dexamethasone for the treatment of relapsed/refractory RAS-pathway mutated paediatric and adult acute lymphoblastic leukaemia (SeluDex): study protocol for an international, parallel-group, dose-finding with expansion phase I/II trial"

### Participant Information Sheet – Adult, Phase I

*To be used only for patients over 16 years of age considering entering dose finding phase*

---

This leaflet provides information about a clinical research trial and is intended to supplement your discussions about the trial with your doctor and nurses. Having read it, you may have further questions and these should be discussed with your consultant or one of the research team.

---

Dear Patient,

We would like to invite you to take part in a non-commercial, clinical trial being run by the University of Birmingham. Before you decide whether to take part in this trial, it is important for you to understand why the research is being done and what it would involve for you.

Please take time to read the following information carefully, and discuss it with your friends and relatives if you wish. Your doctor will go through the information sheet with you, and answer any questions you may have.

- Part 1 tells you the purpose of this trial, and what will happen to you if you take part
- Part 2 gives you more detailed information about the conduct of the trial

If there is anything that is not clear, or if you would like more information, please ask your doctor.

Take your time to decide whether or not you wish to take part. If you decide not to take part, this will not affect the quality of your care.

#### PART 1

##### What is the purpose of the trial?

The purpose of this trial is to test a new drug called selumetinib in combination with another drug called dexamethasone. The trial specifically includes those patients who have a relapse of their acute lymphoblastic leukaemia (ALL) or refractory ALL, and who have an identified change (mutation) in a particular gene in their cancer's DNA. We would like to see what effect combining these two drugs has on you and your leukaemia. This will include looking at how well this treatment works, finding out more information about how it affects the disease, and to see how safe the drugs are.

During Phase I, the part you have been invited to participate in, the trial will look at establishing what is the most suitable dose level of selumetinib in combination with dexamethasone. The purpose of this step of the research is to see what dose of selumetinib we can safely give to participants. We will get some preliminary information regarding the effectiveness of this combined treatment. We will also use results from your tests to see how the treatment affects the body.

After the Phase I part of the trial is complete, further patients who have not taken part in Phase I will be invited to participate in Phase II. The Phase II part of the trial will look at a dose level of selumetinib which has already been established in Phase I as being the most effective in combination with dexamethasone. The purpose of this step of the research is to develop further evidence about this recommended dose, and to see what effects the combination of these medications will have on participants' leukaemia. We will also continue to use results from tests to see how the treatment affects the body.

##### Why have I been invited to participate?

You have been invited to take part in the Phase I part of the trial because you have been diagnosed with relapsed ALL or with refractory ALL. If you have relapsed ALL and you are over 18 years of age this will be at least your first relapse whereas if you are 16-18 years of age this will be at least your second relapse. You will have previously consented to us undertaking a genetic test, or already had this testing done at hospital as part of your normal care, to assess the genetic status of your disease. This test has shown that your leukaemia carries a mutation in the RAS pathway, which means that you are a suitable candidate to take part in the SeluDex trial. Up to 42 participants will be recruited to the trial in total.

##### Do I have to take part?

No, you do not have to take part. It is up to you to decide whether or not to join the trial. Your doctor will describe the trial, and will go through this information sheet with you. They will also give you a copy of the information sheet to keep. If you decide to be part of the trial, you will be asked to sign a consent form to show you have agreed to participate. You are free to withdraw at any time, without giving a reason, and this will not affect the standard of care you receive thereafter.

##### What are the drugs being tested?

The investigational drugs being tested in this trial are called 'selumetinib' and 'dexamethasone'.

Selumetinib is manufactured and supplied by AstraZeneca, and it has not yet been licensed by the Medicines and Healthcare products Regulatory Agency (MHRA) and therefore only available as part of a clinical trial. As researchers learn more about the gene changes in cells that cause cancer, they have been able to develop drugs that target these changes. Selumetinib is an example of such a drug. Cells normally grow in an orderly way. Chemical messages or signals tell them when to grow and when to stop. But if a gene becomes abnormal, a protein called MEK can produce a signal which makes cancer cells grow abnormally. Selumetinib is a MEK inhibitor, which means that it stops MEK proteins from sending signals to cancer cells for them to grow. This may stop or slow down the growth of cancer cells. As of 31 January 2020, approximately 4,335 patients have received selumetinib on its own, or in combination with another drug. You will take one dose of selumetinib on the first day of your first cycle and then from day 4 you will continue taking the drug throughout your treatment.

Dexamethasone is already used for the treatment of a number conditions including cancers and leukaemias, and belongs to a group of drugs known as glucocorticosteroids. These are important in the treatment of leukaemia due their ability to stimulate the death of cancer cells. Dexamethasone will be given as a tablet or oral suspension. You will not start taking dexamethasone until the second day of your first cycle, and will take this drug intermittently throughout treatment. You will take dexamethasone for 3 days during your first week of your first cycle, not take for 3 days and then take for 4 days until the beginning of cycle 2. After this, you will take dexamethasone just for the first five days of each cycle.

The research team at your hospital will provide you with more information on the routine for taking your medicine if you decide to take part in the trial.

If your leukaemia has also been detected in the central nervous system (CNS) (after a test called a lumbar puncture), additional treatment with intrathecal methotrexate (IT MTX) may be administered to help treat the leukaemia in the CNS.

It appears that it is more difficult for ALL participants receiving selumetinib and dexamethasone treatment in the SeluDex trial to ward off infections. Therefore participants have an increased risk of complications from this such as sepsis, which arises when the body's response to infection causes injury to its own tissues and organs requiring quick treatment. This could be due to the way selumetinib works in the body and that dexamethasone can hide symptoms of infection. Because of this you will receive a specific antibiotic (fluoroquinolone) during your first two cycles of treatment and another specific antibiotic (co-trimoxazole) throughout your treatment on the trial to help prevent this. Having preventative antibiotics is often part of your regular cancer care. Your doctor will also discuss with you that if you experience any signs of infection or sepsis (fever, fast heartbeat or breathing) you should contact your treating hospital straight away.

#### What will happen to me if I take part?

If you decide to take part, you will be asked to sign an Informed Consent Form to show that you have agreed to take part in the trial. After you have signed the consent form, you will undergo a period of screening tests to determine whether you are eligible and that it is safe for you to enter the trial.

##### Before you begin the trial:

You will need to have the following screening tests to ensure that it is appropriate and safe for you to take part. Some of these procedures are extra to your regular cancer care. If you have had some of them recently, they may not need to be repeated.

###### Routine tests and procedures

The research team will perform some routine checks. These will include:

- blood, urine tests
- an electrocardiogram (ECG), which will record the electrical activity of the heart
- a review of your medical history

- a physical examination
- assessment of your vital signs including blood pressure, temperature, heart rate
- review of your performance status, which is a measure of your general health and how your disease affects your daily routine
- you will also be asked questions about your health (e.g. date of diagnosis of your conditions, details of previous treatment or any other illnesses you may have) and what medicines you are taking at the moment
- a pregnancy test if you are female and of child-bearing potential

##### Extra tests and procedures

In addition to the tests above, an echocardiogram (ECHO) will be performed. This will record how your heart pumps blood and how the valves inside your heart look and work.

You will also have an eye examination. During the eye exam, the ophthalmologist will examine your eyes with a number of different instruments. They will look at the different structures of your eye to check for abnormalities. They may put drops in your eyes which will temporarily affect your vision, so you shouldn't drive yourself home. This test will be performed at screening only, but may be repeated whilst on treatment if deemed necessary by the trial doctor.

Fluid will be taken by lumbar puncture to test for leukaemia in the CNS which may or may not be routine. During this test treatment with intrathecal methotrexate (IT MTX) may be administered to help prevent or treat CNS relapse. This test will be performed for all participants at screening, and repeated at cycle 1 day 28 for those with CNS positive disease.

Please also refer to the summary table at the end of this information sheet. The screening period may last up to 3 weeks. If you are eligible to take part in the trial, you will be registered and given a unique trial number. If your doctor feels it is appropriate for your medical care, you may be given a course of one of three drugs (either dexamethasone, prednisolone or hydroxycarbamide) during the screening period. This treatment will end either seven days prior or up to one day prior to you registering and starting on the trial depending on the choice of pre-treatment.

##### During the trial:

You will be expected to do the following things during the trial:

- You will have to take selumetinib, twice-daily, continuously throughout the trial, and intermittent dexamethasone during the first and start of the second cycle and then for the first five days of each subsequent cycle
- You will have to have preventative antibiotic treatment
- Whilst taking the trial drugs you will be given a Patient Diary in which to record when you take your medication. Your research nurse will show you how to complete it. The diary will help you to remember if you have taken your dose and to record any side effects. If you accidently miss a dose, it is important that this is recorded in order for your doctor to see what medication you have taken. You will need to bring the diary to your clinic visits, and you will be issued with a new diary at the start of each new cycle of treatment. In addition, you must return the empty bottles and any unused tablets to your doctor when requested
- You will need to come to clinic for scheduled visits as indicated in the table at the end of this leaflet. In this trial, each 28-day period (4 weeks) is called a cycle of treatment
- You will be given a Patient ID Card which provides emergency contact details for the trial team. You should carry this with you at all times and present it to a doctor if you are admitted to a hospital

##### Routine tests and procedures

The research team will perform the following routine tests at some or all of the visits to see how the trial medication is affecting your body (please see the table at the end of this information sheet which shows when each assessment or activity is done): physical exam, pregnancy testing if you are female and of child-bearing potential, vital signs, performance status, ECG, and lab safety tests. You will also be asked questions about your health and what medicines you are taking at the moment.

##### Extra tests and procedures

Additional research blood samples and echocardiograms will be taken at various points during the trial treatment period. A full list of tests and procedures is included at the end of this information sheet.

#### How long will I be in the trial?

Most participants are expected to receive trial treatment for at least 1 or 2 cycles, and initially the number of cycles of trial treatment is six (approximately 6 months). Following this, participants who are considered to be receiving clinical benefit can remain on trial and have continued access to trial treatment whilst the trial is still ongoing after approval from AstraZeneca. If one drug is permanently stopped (for example due to serious side effects), your trial doctor may decide whether it is in your best interest to continue taking the other drug. If the drugs are not helping you, or you are having serious side effects, or you simply do not wish to continue with the trial treatment, you can stop the trial and you can discuss alternative treatment options with your doctor.

#### After you stop taking the trial drug

When you have finished taking the trial medication, you will need to come in to the clinic for an end of treatment visit. You will undergo the same routine and extra tests and procedures as performed during the trial (outlined in the table at the end of this information sheet). These are done to see how the trial affected your health. Depending on how well you responded to treatment and if your hospital has the appropriate facilities you may be asked to give a sample of your bone marrow or blood using a hospital consent form to store for future research. This would enable researchers to obtain the samples in the future to look if the RAS pathway activating mutation that was identified in your leukaemia at the start of the trial is still there to learn more about how effective the treatment is.

In addition, you need to come in for a follow-up visit 28 days after the last trial medication was taken. Any side effects you may still have, and other medication you may be taking will be reviewed.

After the follow up visit at 28 days, you will be regularly reviewed as part of your ongoing care by your usual doctor.

#### Payments

You will not receive any money for taking part in this trial.

#### What will I have to do?

You will be required to:

- Attend the hospital as requested for assessments on your progress. You will need to have regular checks. Any side effects of this medicine may continue after you finish your treatment so it is important to report side effects promptly

- Take selumetinib capsules, commencing with one dose on Day 1 and twice daily from day 4 after pre-treatment assessments are completed. Your research nurse will give you guidance on this. Capsules should be taken whole, on an empty stomach (no food or drink other than water for 2 hours prior to dosing and 1 hour after dosing) and with a glassful of water
- Selumetinib doses should be taken approximately 12 hours apart at the same time points each day
- Take dexamethasone on day 2-4, 8-11, 15-18 and 22-25 of your first treatment cycle, day 1-4 of your second treatment cycle, then during the first five days of each subsequent cycle
- Have preventative antibiotic treatment
- Report any symptoms to the trial doctor when you attend for clinic appointments. In between clinic appointments, call the study team if you experience any severe side effects
- If you are on regular medication or herbal supplements, please make sure your oncology doctor and research nurse know about them before you start your treatment so that they can be checked for compatibility with your trial treatment
- After starting your trial treatment, if any new medication is required, it is important that you tell the doctor prescribing the new medication that you are in a clinical trial and are taking selumetinib and dexamethasone. A concomitant medication and restrictions card is provided to carry with you. It is also important that you tell the oncology doctor if you have been asked to start taking any new medications
- Do not have any vaccinations without your Doctor’s approval
- If you are going to have surgery or any other procedures, tell your doctor or dentist you are taking part in this trial and inform them of the trial medication
- Bring all unused or remaining trial medication with you to each clinic visit
- Complete a diary to record when you took your trial medication, and bring the diary with you to your clinic appointments. The diary will contain more specific instructions about how to take your medication. A new diary will be provided to you at the beginning of each cycle of treatment
- Stop wearing contact lenses if you experience any mild to moderate eye symptoms. Also, you must not use any eye drops for the treatment of eye symptoms (unless agreed by your doctor). You are advised to talk to your doctor if you have any concerns
- Refrain from:
    - Eating or drinking large amounts of grapefruit/grapefruit juice (no more than a small glass of grapefruit juice (120ml) or half a grapefruit daily)
    - Taking any vitamin or herbal supplements
- Avoid excessive sun exposure, and use adequate sunscreen protection if sun exposure is anticipated
- You must not take Vitamin E vitamins or supplements
- It is recommended that you apply alcohol-free moisturiser to your skin before bedtime.

#### What are the possible side effects of the treatment?

You may have side effects while on the trial. All treatments can have side effects. Everyone taking part in the trial will be monitored carefully for any side effects, however doctors don’t know all the side effects that may happen. Side effects may be mild or very serious. Your health care team may give you medicines to help lessen side effects or in some cases treatment may be delayed.

Participants in this trial will be at risk of known and unknown side effects of selumetinib and dexamethasone, as well as potential unknown risks due to the combination of the 2 drugs.

Participants with central nervous system (CNS) positive disease may be treated with intrathecal methotrexate (IT MTX) so may also be at risk of methotrexate induced neurotoxicity.

In the event of severe toxicity, and at the advice of your doctor, trial treatment may be interrupted temporarily until side effects have eased. Following an interruption, trial treatment could be restarted at a lower dose. If the side effects have not sufficiently reduced, trial treatment will be permanently stopped.

##### Selumetinib

Selumetinib is a new drug. More than 3,080 people have had the drug so far, and the drug has been very well tolerated. As it is a new drug, there may be side effects from taking selumetinib that doctors don't know about yet. The side effects already associated with selumetinib are listed below.

###### Common side effects of selumetinib:

More than 10 in every 100 people (10%) have one or more of these:

- Skin rash, which may be itchy in about 7 out of 10 people (70%). The majority of events are mild, but in some cases (around 10%) this can be more serious and require treatment
- Diarrhoea in about 5 out of 10 people (50%). It may occur within days of starting selumetinib treatment, and participants are advised to call the trial team as soon as possible so that treatment can be started promptly to help manage these side effects
- Feeling sick in about 5 out of 10 people (50%), or being sick in about 3 out of 10 people (30%). Participants are advised to call the trial team as soon as possible so that treatment can be started promptly to help manage these side effects
- Feeling tired or weak in around 4 out of 10 people (40%)
- Swelling of the hands or feet occurs in around 3 out of 10 people (30%)
- Shortness of breath in around 2 out of 10 people (20%). If you experience difficulty breathing or shortness of breath, call the trial team immediately
- Swelling of the face or extremities is commonly reported in patients receiving selumetinib
- Patients may also experience soreness or inflammation of the mouth, which may begin within days of starting treatment and usually occurs within the first month of treatment

###### Less common side effects of selumetinib:

Between 1 and 10 in every 100 people (1%-10%) have one or more of these:

- Swelling of the face, hands or feet
- Mouth sores, dry mouth
- High temperature
- Dry skin and soreness or infection in the skin around fingernails or toe nails. Sun exposure should be avoided when receiving selumetinib treatment
- Blurred vision – this has been mild or moderate in all reported previous cases. There is around a 5% chance that this may affect your daily living, such as using the telephone
- An increase in blood pressure
- Changes in the way your liver works
- Small decrease in cardiac (heart) function – this is usually mild
- Changes in your lungs – this will be carefully monitored by your doctor
- A lower than normal amount of blood cells that carry oxygen (anaemia)

Early information from clinical studies suggests that people of Asian origin may experience higher blood levels of selumetinib than non-Asian subjects. Higher levels of selumetinib in the blood may cause more side effects. Your trial doctor will discuss this information with you if it might affect your participation in the trial.

##### Dexamethasone

This drug has a known side effect profile as listed below:

- Increased appetite/weight gain
- Fluid retention
- An increase in blood pressure
- Excessive glucose in the bloodstream (hyperglycaemia)
- Steroid-induced diabetes mellitus
- Increased sensitivity to infection
- Disturbance of sleep patterns
- Low mood/irritability
- Acne type skin rashes/stretch marks
- Inflammation of the pancreas
- Death of bone tissue due to a lack of blood supply (avascular necrosis)

##### Intrathecal Methotrexate (IT MTX)

IT MTX may be given to trial participants in whom the leukaemia was found in the fluid taken by lumbar puncture, these are called "CNS positive participants". This drug has the potential for rare cases of neurotoxicity. Patients can experience acute toxicity (headache, nausea/vomiting, stiff neck) or delayed side effects (seizures, altered levels of consciousness and encephalopathy). It has been used for several decades in combination with intensive chemotherapy for the treatment of paediatric and adult ALL order to prevent CNS relapses. All participants will be regularly assessed for the occurrence of adverse effects, and CNS positive participants will also have regular full neurological examinations.

#### What are the possible benefits of taking part?

We cannot predict whether you will benefit directly from taking part in this trial. All participants in the trial will be closely monitored and supported by the research team. It is possible that your disease may respond to the trial treatment. We cannot guarantee that this treatment will help you, but the information we get from this trial will help us to improve the treatment of people with ALL in the future. You will not be paid for the use of your samples, even if the research does lead to the development of new medical products. Such developments take time and are unlikely to be critically dependent upon samples from any particular participant.

#### What are the possible disadvantages and risks of taking part?

It is possible that this combination of treatment will not work and you will still experience side effects.

##### Blood samples

You may experience a little discomfort when a needle is put into a vein or a port-a-cath to take blood samples. There may be slight pain, a small amount of bleeding, discolouration, or bruising at the site where the needle is inserted (but this will clear after a week or two). There is also a risk of infection or phlebitis (abnormal blood clot), but these rarely happen. Blood samples will be grouped together and taken with routine samples as much as possible to minimise the number of times a needle is inserted into your vein.

##### Bone marrow samples

A bone marrow test is taken during your screening visit and again after 4 weeks of treatment to see how well you have responded to the treatment. This is used to look for the amount of leukaemia that you have left and is done by microscope examination and in addition a minimal residual disease (MRD) test is performed on the screening sample and on future samples for participants who have responded well. The site of the bone marrow examination might be painful for a few days afterwards. A bone marrow examination may be performed under a general anaesthetic.

##### Lumbar puncture

A lumbar puncture is performed during screening and might get repeated in case the leukaemia is found in the fluid taken during the initial lumbar puncture. In this case your physician might want to treat you with IT MTX (see paragraph on side effects of treatment). A lumbar puncture may be performed under a general anaesthetic together with a bone marrow examination. Side effects of the procedure include headaches, local discomfort, bruising, bleeding and a low risk of infection.

##### Possible harm to the unborn child and pregnancy

Selumetinib may have a harmful effect on a baby developing in the womb when administered to a pregnant woman. Reproductive toxicology data indicate that selumetinib has adverse effects on embryo foetal development and survival, at dose levels that do not induce maternal toxicity in animal models. It is therefore essential that both men and women taking the trial treatment must agree to use a reliable form of contraception during the trial, e.g. oral contraceptive and condom, intra-uterine device (IUD) such as Mirena Coil and condom, diaphragm with spermicide and condom. Total abstinence is only an acceptable method of contraception when it is your usual and preferred lifestyle choice. You should continue the use of adequate contraception for at least 12 weeks after the treatment has finished.

##### Information for women

You should not take part in this trial if you are pregnant, breastfeeding or may become pregnant during the trial period. If you are female and of childbearing potential, you will have a pregnancy test during screening and then monthly whilst on trial. This may be repeated if pregnancy is suspected whilst you are on the trial. You must agree to use two reliable forms of contraception (as detailed above) for two weeks prior to entering the trial, during the trial, and for at least 12 weeks after the treatment has finished.

If you do become pregnant during the course of the trial, please tell your trial doctor immediately. The pregnancy will need to be monitored and information about the outcome of your pregnancy will be collected from your medical notes and those of your baby.

##### Information for men

If you have a partner who is pregnant or who could become pregnant you must agree to use two reliable forms of contraception (as detailed above) for two weeks prior to entering the trial, during the trial, and for at least 12 weeks after the treatment has finished.

If your partner becomes pregnant whilst you are on the trial treatment, please tell your doctor immediately. We would also like to collect information about the outcome of the pregnancy. If your partner becomes pregnant during the trial, we will ask your partner to consent to this monitoring, if she is happy to do so.

#### What are the alternatives for treatment?

It is important that you discuss any possible treatment alternatives with your doctors before deciding whether to enter this trial. If you decide not to participate in this trial, you will receive standard or palliative care, or may enter another early phase trial (if available).

#### What happens when the research trial stops?

The trial treatment will not be available after the end of the trial. You will receive trial treatment should it be of benefit only for the duration of the trial. Initially this will be for 6 cycles (approximately 6 months). After this if treatment is considered to be of benefit to you then you may remain on treatment by continuing on the trial whilst it is ongoing following approval from

AstraZeneca. The schedule of assessments for treatment continuation will follow cycles 3-6 (see table at the end of this document). At the end of the research trial, or if you withdraw from the trial before it ends, your trial doctor will assess your symptoms and discuss your options and prescribe further alternative treatment if appropriate. If the academic body sponsoring the research trial decides to stop the trial before it has finished, your trial doctor will explain the reasons why and arrange appropriate care for you.

#### What if there is a problem?

Any complaint about the way you have been treated during the trial or any possible harm you might suffer will be addressed. The detailed information on this is given in part 2 of this leaflet.

#### Will my taking part in the trial be kept confidential?

Yes. We will follow ethical and legal practice, and all information about you will be handled in confidence. The details are included in Part 2 of this leaflet.

This completes Part 1 of the Participant Information Sheet

If the information in Part 1 has interested you, and you are considering participation, please read the additional information in Part 2 before making any decision

#### PART 2

##### What if relevant new information becomes available?

Sometimes during the course of a research trial, new information becomes available about the disease or drug that is being studied. If this happens, your doctor will tell you about it and discuss with you whether you want to continue on the trial. If you decide to withdraw, your doctor will make arrangements for your care to continue in a different way. If you decide to continue in the trial you may be asked to sign an updated consent form. If the trial is stopped for any other reason, we will tell you and arrange your continuing care.

##### What will happen if I don't want to carry on with the trial?

You are free to withdraw from the trial at any time. You do not have to give a reason and your future treatment will not be affected. Your doctor will discuss your treatment options with you and will offer you the most suitable treatment available. However, if you choose to withdraw entirely from the trial, we would still like to use the information we have collected about you up until withdrawal. If you just want to stop trial treatment but are happy to be seen in accordance with the schedule specified in this information leaflet please let your local trial team know. Alternatively we would like to ask your permission for your hospital to continue to send information about your progress to the SeluDex Trial Office. This is information that is routinely taken and you will not need to do anything extra. Any stored blood or tissue samples collected specifically for this trial up until the point of withdrawal will be retained and analysed. With your permission, the SeluDex Trial Office would also like to obtain information about your progress from the cancer registries and national databases (for example NHS Digital).

##### What if there is a problem?

You will be closely monitored both during and after treatment, and any side effects you experience will be treated.

**Complaints**: If you have a concern about any aspect of this trial, you should ask to speak with the trial doctor who will do their best to answer your questions (see contact number at the end of this form). If you remain unhappy and wish to complain formally, you can do this through the NHS Complaints Procedure. Details can be obtained from the hospital.

**Harm**: Every care will be taken in the course of this clinical trial. However, in the unlikely event that you are harmed and this is due to someone's negligence then you may have grounds for legal action for compensation against the trial sponsor (University of Birmingham) or the NHS Trust treating you, but you may have to pay your legal costs. NHS Trust and Non-Trust Hospitals have a duty of care to patients treated, whether or not the patient is taking part in a clinical trial and the normal NHS complaints mechanisms will still be available to you. The sponsor of the trial does not hold insurance against claims for compensation for injury caused by participation in this trial and they cannot offer any indemnity.

Participants may also be able to claim compensation for injury caused by participation in this clinical trial without the need to prove negligence on the part of the sponsor or another party. You should discuss this possibility with your trial doctor in the same way as above.

Regardless of this, if you wish to complain, or have any concerns about any aspect of the way you have been approached or treated by members of staff or about any side effects (adverse events) you may have experienced due to your participation in the clinical trial, the normal NHS complaints mechanisms are available to you. Please ask your trial doctor if you would like more information on this. Details can also be obtained from the Department of Health website: http://www.dh.gov.uk.

#### What will happen to the samples that I give?

Routine safety blood and urine samples will be analysed at your hospital laboratory throughout the course of the trial to see how the trial medication is affecting your body (pharmacodynamic analysis) and how your body processes the trial medication (pharmacokinetic analysis).

Throughout the course of the trial, additional samples will be taken for the purpose of the research. These samples will be labelled and identified with a unique sample ID number, your trial number and sometimes date of birth and initials. Samples will be sent to three different locations:

- Samples collected for pharmacokinetic analysis will be sent to Newcastle University
- Samples for pharmacodynamic analysis will be initially stored at the biobank at the University of Birmingham and and may be subsequently analysed at Newcastle University and at AstraZeneca
- Minimal Residual Disease (MRD) samples, if not tested routinely at your hospital, will be sent to University College, London for analysis.

Any samples remaining at the end of the trial may be stored and used for future ethically approved research which may involve genetic analysis, animal or other laboratory work at commercial or private institutions, and which may take place in the UK or overseas. If your samples are used, they will be anonymised.

#### Will any genetic tests be done?

Prior to being approached by your doctor to consider taking part in this trial, you will have consented to give a sample to assess whether your disease is positive for a RAS pathway activating mutation in its genetic make up.

#### What will happen to the results of the research trial?

At the end of the trial, the information collected will be analysed and published in recognised medical journals. You will not be identified in any report or publication. The results will help doctors to decide how to treat relapsed ALL in the future. Participants taking part in this trial can find out about the results from their trial doctor once the results have been published. The results will also be available on the Cancer Research UK website and the CRCTU website (http://www.birmingham.ac.uk/crctu). We expect the first results to be available in approximately 3 years.

#### Who is organising and funding the research?

The trial is an investigator-initiated and investigator-led trial, and is being carried out by a network of doctors across the UK and internationally. The trial is sponsored by the University of Birmingham and is being coordinated by the Cancer Research UK Clinical Trials Unit (CRCTU). Financial support for the trial is provided by the charity Cancer Research UK and from a pharmaceutical company called AstraZeneca. Your doctor will not receive any payments for talking with you about, or recruiting you into, this research trial.

#### Who has reviewed the trial?

All research in the NHS is looked at by independent group of people called a Research Ethics Committee to protect your safety, rights, wellbeing and dignity. This trial has been reviewed and given favourable opinion by a Research Ethics Committee, and by the local Research and Development department at your hospital's Trust. Whilst the trial is ongoing, the results will be reviewed by a Trial Safety Committee (TSC) to ensure that it is appropriate to continue with the trial.

#### Will my taking part in this trial be kept confidential?

All your details and information collected about you for this research trial will be subject to the General Data Protection Regulation (GDPR) and the Data Protection Act 2018 and will be kept totally confidential. Your initials, date of birth, ethnicity, hospital number and NHS number (or Community Health Index (CHI) in Scotland) will be supplied to the Cancer Research UK Clinical Trials Unit when you are entered in to the trial along with a copy of your consent form. In routine communication, your hospital the Cancer Research UK Clinical Trials Unit will refer to you only by a unique trial number allocated to you, and/or your date of birth and initials. All information about you will be securely stored, in both electronic format and paper form by the researchers, and will only be accessible by authorised personnel.

Occasionally, we may need to check your medical records to make sure that the information provided about you is accurate. This will be done either by clinical staff or by designated trial personnel. It may also be necessary to allow authorised personnel from government regulatory agencies, the sponsor of the trial (the group legally responsible for the conduct of the trial) and/or NHS bodies to have access to information about you. This is for your protection, and is to ensure that the research trial is being conducted to the highest possible standards.

If you agree to take part in this trial, then, in addition to the authorised personnel who review the main trial information, medical information about you may be passed on to researchers for future medical research. The information that they will be given for their work relates to your medical condition and treatment only and will not be directly linked to your identity. Researchers will not be able to contact you directly about their research in the future and you will not be identified in any reports or publications resulting from the study.

*At <insert hospital name> this trial will be managed by the <insert name> Clinical Trials Unit at <insert location> where the clinical team looking after you reside. Therefore it will be necessary for identifiable information to be transferred to and held by the Clinical Trials Unit. This includes a copy of the consent form which you will sign if you agree to take part in the trial. This information will remain confidential at all times.

*Delete as appropriate

The University of Birmingham is the sponsor for this study based in the United Kingdom. We will be using information from you and/or your medical records in order to undertake this study and will act as the data controller for this study. This means that we are responsible for looking after your information and using it properly. The University of Birmingham will keep identifiable information about you for up to 25 years after the study has finished.

Your rights to access, change, or move your information is limited, as we need to manage your information in specific ways in order for the research to be reliable and accurate. If you withdraw from the study, we will keep the information about you that we have already obtained. To safeguard your rights, we will use the minimum personally-identifiable information possible.

You can find out more about how we use your information in our Privacy Policy on our website (www.birmingham.ac.uk/crctu).

The NHS via your hospital(s) will collect information from your medical records for this research study in accordance with our instructions. Your hospital(s) will use your name, NHS number and contact details to contact you about the research study, and make sure that relevant information about the study is recorded for your care, and to oversee the quality of the study.

The NHS will keep identifiable information about you from this study for up to 25 years after the study has finished.

#### Involvement of the General Practitioner/Family Doctor (GP)

As this trial requires you to take a drug which may cause side-effects, it is important that your General Practitioner (GP) is informed that you are taking part in the trial. With your permission your trial doctor will notify your GP that you intend to participate in the trial. In addition, we may ask them to provide information on your progress. If we do need to contact your GP for any follow-up information, we will need to use your full name in our correspondence.

#### Further information and contact details

If you have any further questions or concerns about your disease or this clinical trial please feel free to discuss them with the doctors and nurses looking after you before deciding to take part in the trial or at any time during the trial. Before you make a decision, you may want to discuss the trial with your family and friends and with your family doctor.

You may also find it helpful to contact the following organisations:

**Cancer Research UK**

An information service about cancer and cancer research studies hosted by Cancer Research UK

www.cancerresearchuk.org/about-cancer/

**Macmillan Cancer Support**

An independent patient advisory group

Macmillan Cancer Support, 89 Albert Embankment, London, SE1 7UQ, UK

www.macmillan.org.uk

**Children's Cancer and Leukaemia Group**

A children's cancer charity for those involved in the treatment and care of children with cancer

CCLGroup, Clinical Sciences Building, Leicester Royal Infirmary, Leicester LE2 7LX

http://www.cclg.org.uk/

Please take as much time as you need to make a decision and then let your doctor know what you have decided so that your treatment can be arranged.

If you have any further questions, you are very welcome to contact the trial doctor who is leading this trial, whose contact details are provided below:

**Local Investigator:** ………………………………………………………………...

**Trial Nurse:** ………………………………………………………………...

**Trial Coordinator:** ………………………………………………………………...

**Emergency Contact Number:** ………………………………………………………………...

**Thank you for reading this information sheet. If you decide to take part you will be given a copy of this information sheet and your signed consent form.**

#### Assessments at Each Visit

| | Prior to start of treatment | Cycle 1 | Cycle 2 | Day 1 of each following cycle | End of Treatment | 28 days follow up |
|---|---|---|---|---|---|---|
| **Laboratory Tests** | | | | | | |
| Blood tests | ✓ | ✓ (Days 1, 8, 15, 22 & 28) | ✓ (Days 1 & 15) | ✓ | ✓ | ✓ |
| Urine sample | ✓ | ✓ (Days 1, 8, 15 & 22) | ✓ (Day 1) | ✓ | ✓ | ✓ |
| Pregnancy test [a] | ✓ | | ✓ | ✓ | ✓ | ✓ |
| **Disease Assessment** | | | | | | |
| Bone marrow testing/MRD Sample | ✓ | ✓ (Day 28) | | ✓ (Cycle 4 & 6 only) | ✓ | |
| Lumbar puncture | ✓ | ✓ (Day 28 if CNS positive) | | | | |
| **Treatment** | | | | | | |
| Selumetinib | | ✓ (Day 1 and continuously from day 4) | | | | |
| Dexamethasone | | ✓ (Day 2-4, 8-11, 15-18 & 22-25) | ✓ (Day 1-4) | ✓ (Day 1-5) | | |
| Preventative Antibiotics | | Throughout treatment | | | | |
| **Research Samples** | | | | | | |
| Blood samples | | ✓ (Day 1 & 4) | ✓ (Day 1) | | ✓ | |
| **Other Assessments and Activities** | | | | | | |
| Medical history | ✓ | | | | | |
| Physical examination | ✓ | ✓ (Day 1, 8, 15 & 22) | ✓ (Day 1 & 15) | ✓ | ✓ | ✓ |
| Performance Status | ✓ | ✓ (Day 1, 8, 15 & 22) | ✓ (Day 1) | ✓ | ✓ | ✓ |
| Vital signs (incl. weight) | ✓ | ✓ (Day 1, 8, 15 & 22) | ✓ (Day 1) | ✓ | ✓ | ✓ |
| Growth Chart (Height) – 16-18yrs only | ✓ | | | ✓ (Cycle 4 only) | ✓ | ✓ |
| ECG | ✓ | ✓ (Day 1) | ✓ (Day 1) | ✓ | ✓ | ✓ |
| ECHO | ✓ | ✓ (Day 28) | | Additional tests may be necessary if your doctor is concerned about side effects | | |
| Eye exam | ✓ | Additional tests may be necessary if your doctor is concerned about side effects | | | | |
| Medication/side effects monitoring | ✓ (Done at every clinic visit from screening through to 28-day follow up visit) | | | | | |

[a] Female participants of childbearing potential only
