## Supplementary Appendix 4 for "Selumetinib in combination with dexamethasone for the treatment of relapsed/refractory RAS-pathway mutated paediatric and adult acute lymphoblastic leukaemia (SeluDex): study protocol for an international, parallel-group, dose-finding with expansion phase I/II trial"

SeluDex Trial Schedule of Events

| | **Screening**[1] (Within 14 days of Trial Entry) | **C1/Day 1**[2] | **C1/Day 2** | **C1/Day 4** | **C1/Day 8**[3] (± 2 days) | **C1/Day 15**[3] (± 2 days) | **C1/Day 22**[3] (± 2days) | **C1/Day 28**[4] (- 3 days during Phase II only) | **C2/Day 1**[5] | **C2/Day 15**[3] (± 2 days) | **C3/Day 1**[6] (- 2 days) | **C4/Day 1**[6, 7] (- 2 days) | **C5/Day 1**[6] (- 2 days) | **C6/Day 1**[6, 7] (- 2 days) | **End of Treat-ment Visit**[8] (+ 14 days) | **28 day Follow Up Visit**[9] (+ 2 days) |
|---|---|---|---|---|---|---|---|---|---|---|---|---|---|---|---|---|
| Informed Consent[a] | x | | | | | | | | | | | | | | | |
| RAS-pathway mutation testing[b] | x* | | | | | | | | | | | | | | x | |
| Medical history[c] | x | | | | | | | | | | | | | | | |
| Performance Status (ECOG, Lansky, or Karnofsky scale) | x | x | | | x | x | x | | x (- 1 day) | | x | x | x | x | | x |
| Vital signs[d] | x | x | | | x | x | x | | x (- 1 day) | | x | x | x | x | x | x |
| Weight (kg) | x | x (-3 days) | | | x | x | x | | x (- 1 day) | | x | x | x | x | x | x |
| Body Surface Area | x | x (-3 days) | | | | | | | x (- 1 day) | | x | x | x | x | | |
| Physical examination[e] | x | x | | | x | x | x | | x (- 1 day) | x | x | x | x | x | | x |
| Serum chemistry[f] | x | x (-3 days) | | | x | x | x | | x (- 1 day) | x | x | x | x | x | | x |
| Full blood count & peripheral blood blast count[g] | x | x (-3 days) | | | x | x | x | x | | x | x | x | x | x | x | x |
| ECG[h] | x | x | | | | | | | x (- 1 day) | | x | x | x | x | | x |
| Urinalysis (dipstick) | x | x | | | x | x | x | | x (- 1 day) | | x | x | x | x | | x |
| Height (cm) | x | | | | | | | | | | | | | | | |
| Bone marrow aspirate[i] | x | | | | | | | x | | | | x (- 3 days) | | x (- 3 days) | x | |
| Cytogenetic analysis & Immunophenotyping | x | | | | | | | | | | | | | | | |
| MRD sample[j] | x | | | | | | | x | | | | x (- 3 days) | | x (- 3 days) | x | |
| Lumbar puncture[k] | x | | | | | | | x | | | | | | | | |
| Serological studies for HBV, HCV, HIV | x | | | | | | | | | | | | | | | |
| Pregnancy test[l] | x (-7 days) | | | | | | | | x (-1 day) | | x | x | x | x | x | x |
| ECHO[m] | x | | | | | | | x (- 3 days) | | | | | | | | |
| Ophthalmological exam[n] | x | | | | | | | | | | | | | | | |
| Adverse events | | x | x | x | x | x | x | x | x | x | x | x | x | x | x | x |
| Concomitant medications | x | x | x | x | x | x | x | x | x | x | x | x | x | x | x | x |
| Growth Chart (Group P only)[o] | x | | | | | | | | | | | x | | | x | x |
| Pharmacokinetic samples (blood)[p] | | x | x | x | | | | | x | | | | | | | |

| | **Screening**[1] (Within 14 days of Trial Entry) | **C1/Day 1**[2] | **C1/Day 2** | **C1/Day 4** | **C1/Day 8**[3] (± 2 days) | **C1/Day 15**[3] (± 2 days) | **C1/Day 22**[3] (± 2days) | **C1/Day 28**[4] (- 3 days during Phase II only) | **C2/Day 1**[5] | **C2/Day 15**[3] (± 2 days) | **C3/Day 1**[6] (- 2 days) | **C4/Day 1**[6, 7] (- 2 days) | **C5/Day 1**[6] (- 2 days) | **C6/Day 1**[6, 7] (- 2 days) | **End of Treat-ment Visit**[8] (+ 14 days) | **28 day Follow Up Visit**[9] (+ 2 days) |
|---|---|---|---|---|---|---|---|---|---|---|---|---|---|---|---|---|
| Pharmacodynamic samples - RNA & Fixed (blood)[q] | | x | x | x | | | | | | | | | | | x | |
| Patient Diary (dispense and review as appropriate)[r] | | x | | | | | | | x | | x | x | x | x | | |
| Selumetinib dispensing | | x | | | | | | | x | | x | x | x | x | | |
| Dexamethasone dispensing | | | x | | | | | | x | | x | x | x | x | | |
| Fluoroquinolone prophylaxis dispensing | | x | | | | | | | x | | | | | | | |
| Co-trimoxazole prophylaxis dispensing | | x | | | | | | | x | | x | x | x | x | | |

Initially trial treatment is for 6 cycles (approximately 6 months) however following this participants who are considered to be receiving clinical benefit can remain on trial and have continued access to selumetinib whilst the trial is open following approval from AstraZeneca. The schedule of assessments for treatment continuation should follow cycles 3-6.

[1] Any assessments done as standard care do not require informed consent and may be provided as screening data if conducted within 14 days of trial entry unless otherwise stated

[2] Weight, Body Surface Area, Serum Chemistry, FBC and peripheral blood blast count may be performed within the 3 days prior to cycle 1 day 1

[3] Visit may occur within the 2 days prior to or after the scheduled visit date

[4] During Phase I bone marrow aspirate and all associated assessments at this visit must occur on cycle 1 day 28 of treatment except for the ECHO which may be performed within the 3 days prior to the scheduled visit date. During Phase II this visit and all associated assessments may occur within the 3 days prior to the scheduled visit date

[5] Performance status, vital signs, weight, BSA, physical examination, serum chemistry, FBC, ECG and urinalysis (dipstick) may be performed within the 1 day prior to the scheduled visit date

[6] Visit may occur within the 2 days prior to the scheduled visit date

[7] Bone marrow aspirate and MRD sample may be performed within the 3 days prior to the scheduled visit date

[8] The End of Treatment assessment visit should be performed within 14 days of the patient's last administration of selumetinib. Should the patient discontinue following a recent bone marrow aspirate visit then a repeat bone marrow aspirate for end of treatment is not required at the discretion of the investigator

[9] Patients will be followed up for 28 days after the last dose of selumetinib, with a clinic visit taking place on day 28. Visit may occur within the 2 days after the scheduled visit date

* RAS-pathway mutation result date does not have to be within 14 days of trial entry but must be confirmed using a sample from the current relapse/disease

[a] Written informed consent must be received before any trial specific procedures occur

[b] Sample from current relapse/disease must confirm presence of a RAS-pathway activating mutation - NRAS, KRAS, FLT3, PTPN11, cCBL, NF1, BRAF, IKZF2, IKZF3, IL7Rα or JAK1. Results from trial office approved local molecular testing are accepted for all of these mutations. Testing for NRAS, KRAS, FLT3, PTPN11 and cCBL mutations can be done centrally by Northern Genetics Services (Previously NewGene Limited) in Newcastle, UK – refer to the laboratory manual for further information. Optional end of treatment sample to be collected from non-responders and stored at sites with local biobank facilities and appropriate ethical approval using local consent to assess clonal evolution of RAS-pathway wildtype ALL

[c] Medical history – including demographics, prior treatment, allergy history, discuss contraception

[d] Blood pressure, pulse measurement, temperature, and respiratory rate to be performed with the patient sitting for 5 minutes prior to the evaluation

[e] Physical examination includes complete review of systems, physical examination of pertinent organ systems, and neurological exam

[f] Includes: Glucose, LDH and creatine kinase (CK) at screening only. Creatinine, Sodium, Potassium, Total Calcium, Total Protein, Phosphate, Magnesium, Urea, Total Bilirubin, AST or ALT ALP and albumin at all timepoints

[g] Full Blood Count with differential count and peripheral blood blast count as an additional request test at all timepoints

[h] Repeat ECG as clinically indicated

[i] Bone marrow aspirate at screening (peripheral blood can be used in the case of a dry tap if peripheral WCC > $50x10^9$/L – screening only); cycle 1 day 28, cycle 4 day 1 and cycle 6 day 1. For any patients who continue treatment after cycle 6, a bone marrow aspirate should be performed every other cycle.

[j] An MRD sample will be collected from the bone marrow aspirate at screening for all patients. At subsequent timepoints an MRD sample should be collected each time a BM aspirate is performed for patients who have achieved at least a CRi and where a marker was identified at screening. Group A: MRD samples will be sent to UCL for analysis. Group P: MRD will be analysed as part of standard of care

[k] A Lumbar puncture to assess CNS status should be done at Screening for all patients and at cycle 1 day 28 for CNS positive patients only

[l] Women of childbearing potential will require a negative pregnancy test (serum or urine) prior to registration and within seven days prior to study drug administration, then monthly

[m] An ECHO should be performed at screening. Repeat on cycle 1 day 28 (or within the 3 days prior) and as clinically indicated. (Group A: LVEF, LVEDV & LVESV, Group P: SF)

[n] Repeat as clinically indicated

[o] Group P only. Body height (standing or lying length where age-appropriate) will be measured. Where possible, a stadiometer should be used. Where this may be difficult in particularly young children, standard measurement methods may be used.

[p] Pharmacokinetic samples at the following time points:

- Cycle 1 day 1: pre first selumetinib dose, then at 30 minutes, 1 hour, 2 hours, 4 hours, 6 hours and 24 hours after the first dose (pre cycle 1 day 2 dexamethasone dose)
- Cycle 1 day 4: pre first selumetinib/dexamethasone dose, then at 30 minutes, 1 hour, 2 hours, 4 hours, 6 hours after the first dose
- Cycle 2 day 1: pre first selumetinib/dexamethasone dose, then at 30 minutes, 1 hour, 2 hours, 4 hours, 6 hours after the first dose

[q] Pharmacodynamic blood samples at the following time points (only if WCC >$10x10^9$/l). PD-RNA samples - all sites. PD-Fixed samples – if facilities available at site as agreed at initiation:

- Cycle 1 day 1: pre first selumetinib dose, then at 1 hour, 4 hours, 6 hours and 24 hours after the first dose
- Cycle 1 day 4: pre first selumetinib/dexamethasone dose, then at 1 hour, 4 hours, 6 hours after the first dose
- End of Treatment visit (one sample only)

[r] All patients/parents/legal guardians will be required to complete a diary, which must be returned to the clinic for checking at each visit. The research nurse should complete the relevant sections including recording the AM and PM doses and the dates they are to be taken on the diary. Patients/parents/legal guardians should be instructed to record daily time of administration of the study drugs in the diary. If a dose is missed, the reason must be noted in the diary by the patient/parent/legal guardian. The patient diary will then be reviewed by the research nurse at the end of each cycle. Patients/parents/legal guardians should be advised to return any unused IMP in the original bottles, in addition to returning any empty bottles.
